# Prevalence and Clinical Significance of Commonly Diagnosed Genetic Disorders in Preterm Infants

**DOI:** 10.1101/2023.07.14.23292662

**Authors:** Selin S. Everett, Miles Bomback, Rakesh Sahni, Ronald J. Wapner, Veeral N. Tolia, Reese H. Clark, Alex Lyford, Thomas Hays

## Abstract

**Background and Objectives:** Preterm infants (<34 weeks’ gestation) experience high rates of morbidity and mortality before hospital discharge. Genetic disorders substantially contribute to morbidity and mortality in related populations. The prevalence and clinical impact of genetic disorders is unknown in this population. We sought to determine the prevalence of commonly diagnosed genetic disorders in preterm infants, and to determine the association of disorders with morbidity and mortality.

**Methods:** This was a retrospective multicenter cohort study of infants born from 23 to 33 weeks’ gestation between 2000 and 2020. Genetic disorders were abstracted from diagnoses present in electronic health records. We excluded infants transferred from or to other health care facilities prior to discharge or death when analyzing clinical outcomes. We determined the adjusted odds of pre-discharge morbidity or mortality after adjusting for known risk factors.

**Results:** Of 320,582 infants, 4196 (1.3%) had genetic disorders. Infants with trisomy 13, 18, 21, or cystic fibrosis had greater adjusted odds of severe morbidity or mortality. Of the 17,427 infants who died, 566 (3.2%) had genetic disorders. Of the 65,968 infants with a severe morbidity, 1319 (2.0%) had genetic disorders.

Conclusions

Genetic disorders are prevalent in preterm infants, especially those with life-threatening morbidities. Clinicians should consider genetic testing for preterm infants with severe morbidity and maintain a higher index of suspicion for life-threatening morbidities in preterm infants with genetic disorders. Prospective genomic research is needed to clarify the prevalence of genetic disorders in this population, and the contribution of genetic disorders to preterm birth and subsequent morbidity and mortality.

**Article Summary:** Genetic disorders were found in 1.3% of preterm infants and at a higher rate (2.0%) in infants who died or developed severe morbidity.

**What’s Known on This Subject:** Previous research described the prevalence and associated short-term morbidity and mortality of trisomy 13, 18, and 21 in preterm infants. The prevalence of other commonly diagnosed genetic disorders and associated short-term morbidity and mortality in preterm infants is unknown.

**What This Study Adds:** In a multicenter, retrospective cohort of 320,582 preterm (<34 weeks’ gestation) infants, we found that 1.3% had genetic disorders diagnosed through standard care. Multiple disorders were associated with increased adjusted odds of morbidities or mortality prior to hospital discharge.

**Contributors Statement Page:** Selin S. Everett conceptualized and designed the study, conducted analyses, drafted the initial manuscript, and critically reviewed and revised the manuscript.

Dr. Thomas Hays conceptualized and designed the study, drafted the initial manuscript, and critically reviewed and revised the manuscript.

Miles Bomback conceptualized and designed the study and critically reviewed and revised the manuscript.

Drs. Veeral N. Tolia and Reese H. Clark coordinated and supervised data collection and critically reviewed and revised the manuscript.

Dr. Rakesh Sahni conceptualized and designed the study and critically reviewed and revised the manuscript.

Dr. Alex Lyford conducted analyses and critically reviewed and revised the manuscript. Dr. Ronald J. Wapner reviewed and critically revised the manuscript.

All authors approved the final manuscript as submitted and agree to be accountable for all aspects of the work.

## Introduction

Preterm infants, particularly those born before 34 weeks’ gestation, experience high rates of severe morbidity and mortality prior to discharge.^1–3^ Multiple factors have been shown to contribute to these outcomes including gestational age, birth weight, and social determinants and health.^1–3^ Genetic disorders have been shown to strongly contribute to morbidity and mortality in related populations including full-term infants and children with critical illness.^4–7^ Genetic disorders are particularly prevalent in stillbirth and fetal disorders.^8–10^ Despite this, there are limited descriptions of the prevalence of genetic disorders in preterm infants. Boghossian et al. described the prevalence, morbidity, and mortality of very low birthweight infants with common aneuploidies.^11,12^ However, no studies to our knowledge have provided a comprehensive description of genetic disorders diagnosed in this population. We therefore sought to achieve the following objectives: 1) determine the prevalence of commonly diagnosed genetic disorders in preterm infants, and 2) determine the association of these genetic disorders with severe morbidity and mortality in preterm infants.

We recently utilized the Pediatrix Clinical Data Warehouse (CDW) in related studies, to investigate the association of congenital renal anomalies with genetic disorders in preterm infants.^13^ CDW contains detailed records of 20% of preterm infants born in the United States. Given this utility and broad representation, we sought to employ a similar strategy in this study.

## Methods

This study was approved by the institutional review board of Columbia University. This report followed the guidelines for cohort studies as outlined by Strengthening the Reporting of Observational Studies in Epidemiology (STROBE). This was a retrospective study using the CDW multicenter cohort of infants born from 23 to 34 weeks’ gestation.

### Prevalence of commonly diagnosed genetic disorders in preterm infants

Multiple genetic disorders, including common aneuploidies, are associated with fetal growth restriction. Therefore, we used a gestational age-based cutoff rather than weight-based. We extracted records from 409,704 infants discharged from 2000 to 2020. We excluded infants with missing covariate data, as well as those transferred after birth (often because of complex illness) and infants transferred prior to discharge (in which the clinical outcome is unknown) to limit ascertainment biases.^15^ After these exclusions, 320,582 preterm infants born < 34 weeks’ gestation remained. Data from NICU admission to discharge were collected, including clinician-noted diagnosis of genetic disorders. This cohort did not undergo prospective, genome-wide genetic testing. Genetic diagnoses were limited to those made during standard care.

### Association of commonly diagnosed genetic disorders with severe morbidity and mortality in preterm infants

We collected infant characteristics (genetic diagnoses, gestational age, normalized birth weight and categorization by Fenton criteria at 10^th^ and 90^th^ percentiles for gestational age and sex,^14^ maternal self-identified race, sex, mode of delivery, exposure to antenatal betamethasone, intubation in the first 72 hours of life, year of discharge, and the presence of congenital anomalies) and data regarding clinical outcomes (diagnoses of severe co-morbidities, mortality, and disposition). Severe morbidities were defined as the following: acute kidney injury (AKI) defined by the diagnosis of oliguria or anuria (ICD-9 788.5 or ICD-10: R34); severe intracranial hemorrhage (ICH) defined as grade III or IV intraventricular hemorrhage by Papile criteria or cystic periventricular leukomalacia;6 medically or surgically treated necrotizing enterocolitis (NEC); severe bronchopulmonary dysplasia (BPD) defined as invasive mechanical ventilation at 36 weeks’ postmenstrual age; severe retinopathy of prematurity (ROP) defined as need for any medical or surgical intervention; culture-positive sepsis defined by any positive blood or urine culture; or shock defined as administration of any vasopressor or inotrope.

We used logistic regression to model the relationship between genetic disorders and severe morbidity or mortality. The following were covariates: year of discharge, gestational age, birth weight (here as a Z-score normalized to gestational age and sex to account for continuous rather than categorical effects), sex, maternal race, mode of delivery, intubation in the first 72 hours of life, presence of a congenital anomaly, and exposure to antenatal betamethasone. We determined crude and adjusted odds ratios (OR) after controlling for these covariates. OR were reported with 99.97% confidence intervals (determined as 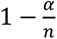) to correct for *n* comparisons of 9 clinical outcomes in 22 categories of genetic disorders and accepting statistical significance at a Bonferroni corrected, two-tailed α of 0.05. Analyses were made using RStudio (version 2022.02.0).^15^

## Results

### Prevalence of commonly diagnosed genetic disorders in preterm infants

Genetic disorders were identified in 4,196 (1.31 %) of the 320,582 infants in this cohort that had data for each of our covariates (**Table 1**). Of the 4,196 infants with a genetic diagnosis, 1927 (45.92%) were females and 2269 (54.08%) were males. The disorders included in the dataset were comprised of 19 specific diagnoses, as well as unspecified aneuploidy, unspecified copy number variant, and infants found to have multiple genetic disorders. Aneuploidies (trisomy 13, 18, 21, and 22, Klinefelter syndrome, Turner syndrome) were present in 1,863 infants, with trisomy 21 accounting for more than half (973) of these cases. Copy number variants (13q deletion, Cri-du-chat syndrome, DiGeorge syndrome) were present in 87 infants. Unspecified aneuploidy and copy number variants were noted in 548 and 35 infants respectively. Eleven distinct monogenic disorders were identified in 2,246 infants. Most (1,986) of these had hematologic disorders. Thalassemia accounted for 1,174 cases. Sickle cell disease was identified in 194 infants and cystic fibrosis in 165. Thirteen infants with multiple genetic disorders were identified. We did not detect differences in the prevalence of classes of genetic disorders diagnosed over time in this cohort (**Figure 1**).

**Figure 1.**
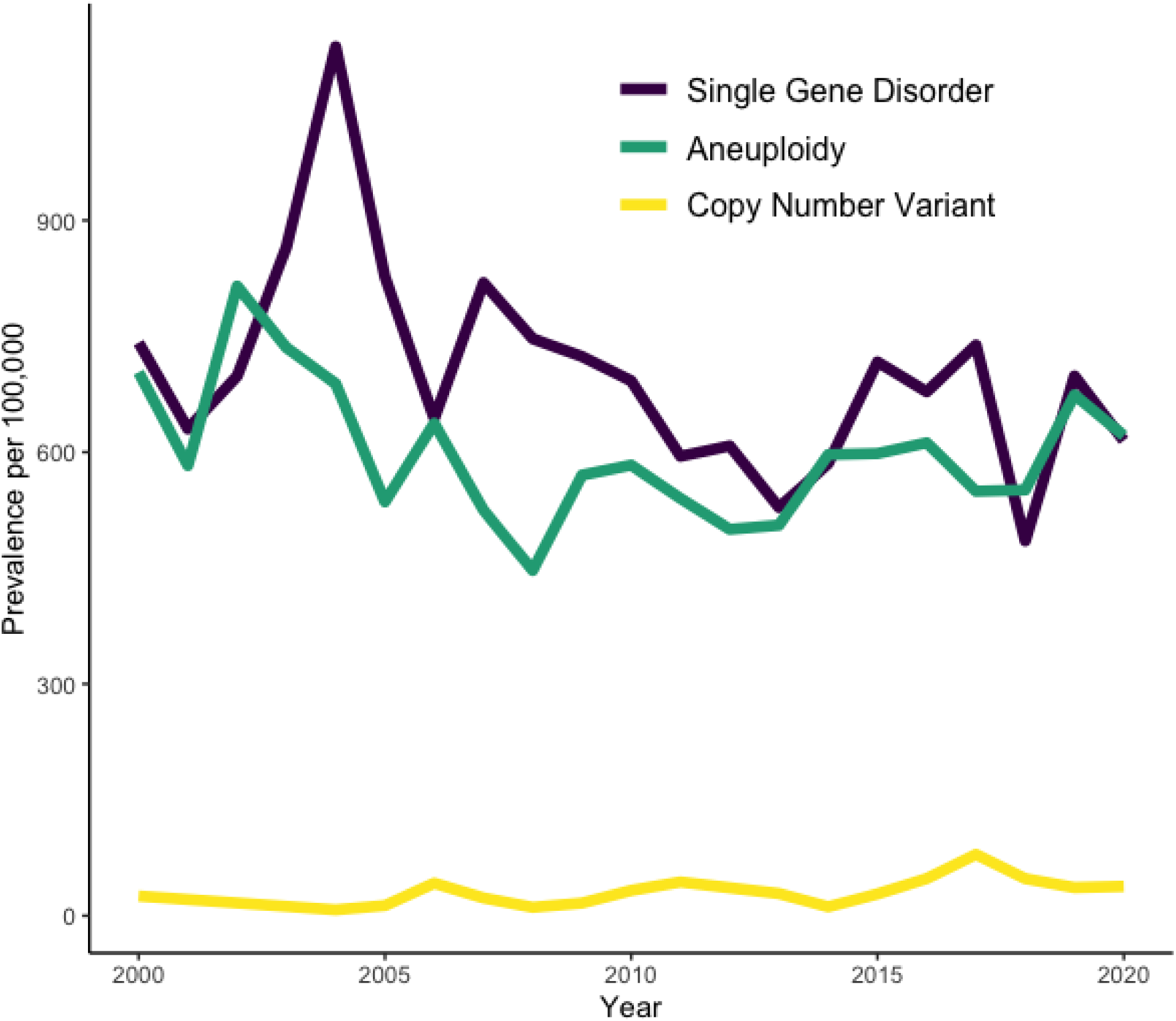
Prevalence of diagnosed genetic disorders over time. We determined the prevalence of the three broad classes (single gene disorder, aneuploidy, and copy number variants) of disorders within preterm infants in this cohort. We did not detect differences in these prevalence rates over time.

**Table 1.** Prevalence of genetic disorders in preterm infants and associated rates of mortality.

| Genetic Disorder | n | n |
| --- | --- | --- |
|  | (Prevalence per 100,000 Preterm Infants) | (Mortality Rate %) |
| Aneuploidy | 1863 (581.1) | 480/1863 (25.8) |
| Trisomy 13 | 110 (34.3) | 93/110 (84.5) |
| Trisomy 18 | 155 (48.3) | 130/155 (83.9) |
| Trisomy 21 | 973 (303.5) | 143/973 (14.7) |
| Trisomy 22 | 1 (0.3) | 1/1 (100) |
| Klinefelter syndrome | 35 (10.9) | 2/35 (5.7) |
| Turner syndrome | 41 (12.8) | 5/41 (12.2) |
| Unspecified Aneuploidy | 548 (170.9) | 106/548 (19.3) |
| Copy number variation disorder | 87 (27.1) | 14/87 (16.1) |
| 13q deletion syndrome | 7 (2.2) | 1/7 (14.3) |
| Cri-du-chat syndrome | 16 (5) | 2/16 (12.5) |
| DiGeorge syndrome | 29 (9) | 4/29 (13.8) |
| Unspecified CNV | 35 (10.9) | 7/35 (20) |
| Monogenic disorder | 2233 (696.5) | 71/2233 (3.2) |
| Hematologic disorder | 1973 (615.4) | 39/1973 (2) |
| Thalassemia | 1174 (366.2) | 26/1174 (2.2) |
| Hemoglobin bart's | 384 (119.8) | 4/384 (1) |
| Sickle cell disease | 194 (60.5) | 5/194 (2.6) |
| G6PD deficiency | 206 (64.3) | 2/206 (1) |
| Hemophilia A | 10 (3.1) | 2/10 (20) |
| Hemophilia B | 5 (1.6) | 0/5 (0) |
| Miscellaneous | 260 (81.1) | 32/260 (12.3) |
| Cystic fibrosis | 165 (51.5) | 16/165 (9.7) |
| Fragile X syndrome | 7 (2.2) | 0/7 (0) |
| Hypophosphatasia | 86 (26.8) | 15/86 (17.4) |
| Fanconi Anemia | 2 (0.6) | 1/2 (50) |
| Multiple Disorders | 13 (4.1) | 1/13 (7.7) |

### Association of commonly diagnosed genetic disorders with severe morbidity and mortality in preterm infants

Following exclusions, 320,582 preterm infants remained for analysis of clinical outcomes. Genetic disorders were prevalent in 4,196 (1.3%) of these individuals. The rate of mortality in preterm infants without genetic disorders in this cohort, and following exclusions was 5.3%. Genetic disorders were present in 566 (3.2%) of preterm infants who died, in 1,319 (2.0%) of preterm infants who experienced any severe morbidity or mortality, in 3,630 (1.2%) of preterm infants who survived to discharge, and in 2,664 (1.1%) of preterm infants who survived to discharge free of severe morbidity. The mortality rate after exclusions for each genetic disorder is provided in Table 1. The highest crude rates of mortality were found in cases of aneuploidy. 84.5 and 83.9% of preterm infants with trisomy 13 and 18 died before discharge. 14.7% of preterm infants with trisomy 21 died before discharge. Mortality rates for preterm infants with copy number variants varied from 12.5% in Cri-du-chat syndrome to 20.0% in cases of unspecified copy number variants. Preterm infants with hematologic disorders had lower observed rates of mortality, with the highest rates found in cases of sickle cell disease (2.6%) and thalassemia (2.2%). Varying rates of mortality were noted in preterm infants with miscellaneous monogenic disorders, including 9.7% mortality for infants with cystic fibrosis.

We then determined the crude and adjusted odds of severe morbidity and mortality for preterm infants with diagnosed genetic disorders as compared to preterm infants without known genetic disorders. The baseline characteristics for covariates used in these analyses are provided for preterm infants with aneuploidies (**Supplemental Table 1**), copy number variants (**Supplemental Table 2**), hematologic disorders (**Supplemental Table 3)**, and miscellaneous monogenic disorders (**Supplemental Table 4**). The absolute numbers of preterm infants experiencing each morbidity and mortality for each diagnosed genetic disorder were also determined (**Supplemental Table 5).**

Preterm infants with aneuploidies had significantly greater adjusted odds of several outcomes (**Table 2**). Preterm infants with trisomy 13, 18, and 21 had greater adjusted odds of death. Preterm infants with trisomy 13 also had greater adjusted odds of AKI. Preterm infants with trisomy 21 had greater adjusted odds of AKI, BPD, sepsis, shock, and death. Preterm infants with copy number variants and hematologic disorders were not found to have significantly greater adjusted odds of morbidity or mortality (**Tables 3 and 4**). The only monogenic disorder diagnosis associated with significantly increased adjusted odds of morbidity or mortality was cystic fibrosis **(Table 5)**. Preterm infants with cystic fibrosis had significantly increased adjusted odds of NEC, BPD, and ROP. Elevated risk of morbidities in preterm infants with hypophosphatasia did not persist after adjustment for known risk factors.

**Table 2.** Odds of severe morbidity or mortality in preterm infants with aneuploidies^1^.

| Outcome | Trisomy 13 (n=110) |  | Trisomy 18 (n=155) |  | Trisomy 21 (n=973) |  | Klinefelter syndrome (n=35) |  | Turner syndrome (n=41) |  |
| --- | --- | --- | --- | --- | --- | --- | --- | --- | --- | --- |
|  | Crude OR | Adjusted OR | Crude OR | Adjusted OR | Crude OR | Adjusted OR | Crude OR | Adjusted OR | Crude OR | Adjusted OR |
| **Mortality or any severe morbidity** | 53.42<br>(15.61, 356.05)* | 61.87<br>(16.15,437.72)* | 29.47<br>(12.34, 90.49) | 28.09<br>(10.56,93.09)* | 2.04<br>(1.58, 2.62) | 2.39<br>(1.75,3.26)* | 1.66<br>(0.35, 6.18) | 0.98<br>(0.16,4.75) | 1.88<br>(0.48, 6.27) | 2.43<br>(0.47,10.51) |
| **Mortality** | 97.18<br>(39.3, 303.91)* | 265.24<br>(93.12,916.46)* | 92.37<br>(43.1, 231.52) | 196.84<br>(81.82,539.66)* | 3.06<br>(2.14, 4.26) | 5.93<br>(3.83,8.95)* | 1.08<br>(0.01, 8.2) | 0.81<br>(0.01,8.14) | 2.47<br>(0.22, 11.21) | 4.17<br>(0.3,29.12) |
| **AKI** | 6.67<br>(2.19, 16.27)* | 4.17<br>(1.24,11.54)* | 3.12<br>(0.84, 8.17) | 1.91 (0.5,5.34) | 3.39<br>(2.15, 5.07) | 4.02<br>(2.43,6.35)* | 2.06<br>(0.02, 15.82) | 1.47<br>(0.01,13.72) | 4.73<br>(0.43, 21.61) | 6.91<br>(0.55,40.06) |
| **ICH** | — | — | 0.32 (0, 2.17) | 0.28 (0,1.99) | 1.09<br>(0.56, 1.9) | 1.29<br>(0.64,2.36) | 1.5<br>(0.01, 11.48) | 1.12<br>(0.01,10.61) | 1.95<br>(0.06, 11.49) | 2.2<br>(0.07,17.02) |
| **NEC** | 1.33<br>(0.12, 5.36) | 0.77 (0.07,3.31) | 0.93<br>(0.09, 3.68) | 0.58 (0.05,2.36) | 1.55<br>(0.85, 2.57) | 1.44<br>(0.78,2.45) | — | — | 1.43<br>(0.01, 10.69) | 1.55<br>(0.01,12.75) |
| **BPD** | 5.37 (0.8, 18.46) | 1.84 (0.26,7.04) | 5.45 (1.19, 15.76) | 1.78 (0.37,5.5) | 4.1 (2.21, 6.91) | 3.32 (1.71,5.94)* | — | — | — | — |
| **ROP** | — | — | — | — | 0.34 (0.02, 1.44) | 0.47 (0.03,2.24) | — | — | — | — |
| **Sepsis** | 0.89 (0.17, 2.73) | 0.49 (0.09,1.65) | 0.76 (0.18, 2.11) | 0.44 (0.1,1.28) | 1.49 (1.02, 2.11) | 1.56 (1.04,2.29)* | 2.95 (0.47, 11.77) | 2.31 (0.34,10.54) | 2.8 (0.51, 10.24) | 3.73 (0.6,16.07) |
| **Shock** | 2.11 (0.74, 5) | 1.08 (0.34,2.89) | 2.38 (1.04, 4.85) | 1.44 (0.58,3.25) | 1.8 (1.28, 2.48) | 2.49 (1.64,3.7)* | 1.23 (0.08, 6.41) | 0.89 (0.05,6.22) | 1.63 (0.19, 6.86) | 2.03 (0.2,11.97) |

Abbreviations: AKI (acute kidney injury), BPD (bronchopulmonary dysplasia), ICH (intracerebral hemorrhage), OR (odds ratio) NEC (necrotizing enterocolitis), ROP (retinopathy of prematurity)
<sup>1</sup>odd ratio (99.97% confidence interval), \* indicates two-tailed, Bonferroni-corrected p-value < 0.05

**Table 3.** Odds of severe morbidity or mortality in preterm infants with copy number variants^1^.

| Outcome | DiGeorge syndrome (n=29) |  | Cri-du-chat syndrome (n=16) |  |
| --- | --- | --- | --- | --- |
|  | Crude OR | Adjusted OR | Crude OR | Adjusted OR |
| **Mortality or any severe morbidity** | 2.56 (0.54, 10.68) | 2.02 (0.29,12.01) | 1.65 (0.13, 11.54) | 1.64 (0.09,17.42) |
| **Mortality** | 2.84 (0.17, 15.59) | 2.59 (0.12,23.83) | 2.54 (0.02, 24.38) | 4.6 (0.04,73.4) |
| **AKI** | — | — | 4.87 (0.05, 47.1) | 5.67 (0.05,79.46) |
| **ICH** | — | — | — | — |
| **NEC** | — | — | 3.99 (0.04, 38.63) | 3.93 (0.04,44.41) |
| **BPD** | 5.85 (0.06, 46.55) | 2.62 (0.02,32.83) | — | — |
| **ROP** | — | — | — | — |
| **Sepsis** | 2.6 (0.3, 12.16) | 2.22 (0.22,12.98) | 3.32 (0.19, 24.16) | 3.91 (0.19,35.63) |
| **Shock** | 1.52 (0.09, 8.35) | 0.98 (0.05,8.17) |  |  |

Abbreviations: AKI (acute kidney injury), BPD (bronchopulmonary dysplasia), ICH (intracerebral hemorrhage), OR (odds ratio) NEC (necrotizing enterocolitis), ROP (retinopathy of prematurity)
<sup>1</sup>odd ratio (99.97% confidence interval), \* indicates two-tailed, Bonferroni-corrected p-value < 0.05

**Table 4.** Odds of severe morbidity or mortality in preterm infants with hematologic disorders^1^.

| **Outcome** | **Thalassemia (n=1174)** |  | **Hemoglobin Bart's (n=384)** |  | **Sickle cell disease (n=194)** |  | **G6PD deficiency (n=206)** |  |
| --- | --- | --- | --- | --- | --- | --- | --- | --- |
|  | **Crude OR** | **Adjusted OR** | **Crude OR** | **Adjusted OR** | **Crude OR** | **Adjusted OR** | **Crude OR** | **Adjusted OR** |
| **Mortality or severe morbidity** | 1.28 (0.99, 1.64) | 0.83 (0.6,1.14) | 0.75 (0.44, 1.23) | 0.86 (0.45,1.57) | 0.91 (0.44, 1.73) | 0.64 (0.26,1.45) | 0.72 (0.33, 1.39) | 0.78 (0.31,1.77) |
| **Mortality** | 0.4 (0.17, 0.79)* | 0.24 (0.1,0.49)* | 0.19 (0.01, 0.8)* | 0.21 (0.01,1) | 0.47 (0.04, 1.83) | 0.32 (0.03,1.47) | — | — |
| **AKI** | 1.42 (0.76, 2.4) | 1.04 (0.54,1.82) | 0.36 (0.02, 1.55) | 0.4 (0.03,1.86) | 1.66 (0.33, 4.94) | 1.4 (0.26,4.64) | 0.85 (0.08, 3.31) | 1.05 (0.1,4.5) |
| **ICH** | 1.36 (0.79, 2.16) | 1.07 (0.61,1.77) | 0.53 (0.09, 1.61) | 0.63 (0.11,2.09) | 0.79 (0.1, 2.82) | 0.67 (0.08,2.64) | 0.87 (0.13, 2.89) | 1.09 (0.16,4.04) |
| **NEC** | 1.37 (0.78, 2.24) | 0.98 (0.54,1.62) | 0.67 (0.13, 1.95) | 0.71 (0.14,2.13) | 0.29 (0, 1.94) | 0.23 (0,1.59) | 1.43 (0.31, 4.06) | 1.54 (0.33,4.58) |
| **BPD** | 1.37 (0.51, 2.92) | 0.99 (0.36,2.19) | 1.04 (0.1, 4) | 1.26 (0.12,5.23) | 2.09 (0.2, 8.18) | 1.82 (0.17,7.81) | 1.96 (0.19, 7.67) | 2.58 (0.24,10.96) |
| **ROP** | 1.77 (0.74, 3.53) | 1.42 (0.56,3.02) | — | — | 0.85 (0.01, 5.65) | 1.04 (0.01,8.08) | — | — |
| **Sepsis** | 1.53 (1.09, 2.09) | 1.14 (0.78,1.61) | 1 (0.48, 1.85) | 1.18 (0.53,2.31) | 1.21 (0.45, 2.66) | 1.02 (0.36,2.46) | 0.78 (0.24, 1.91) | 0.84 (0.24,2.2) |
| **Shock** | 1.13 (0.78,<br>1.6) | 0.75<br>(0.49,1.11) | 0.66 (0.27,<br>1.34) | 0.86<br>(0.32,1.96) | 0.63 (0.16,<br>1.66) | 0.44<br>(0.1,1.34) | 0.43 (0.09,<br>1.29) | 0.54<br>(0.1,1.87) |

Abbreviations: AKI (acute kidney injury), BPD (bronchopulmonary dysplasia), ICH (intracerebral hemorrhage), OR (odds ratio) NEC (necrotizing enterocolitis), ROP (retinopathy of prematurity)
<sup>1</sup>odd ratio (99.97% confidence interval), \* indicates two-tailed, Bonferroni-corrected p-value < 0.05

**Table 5.**
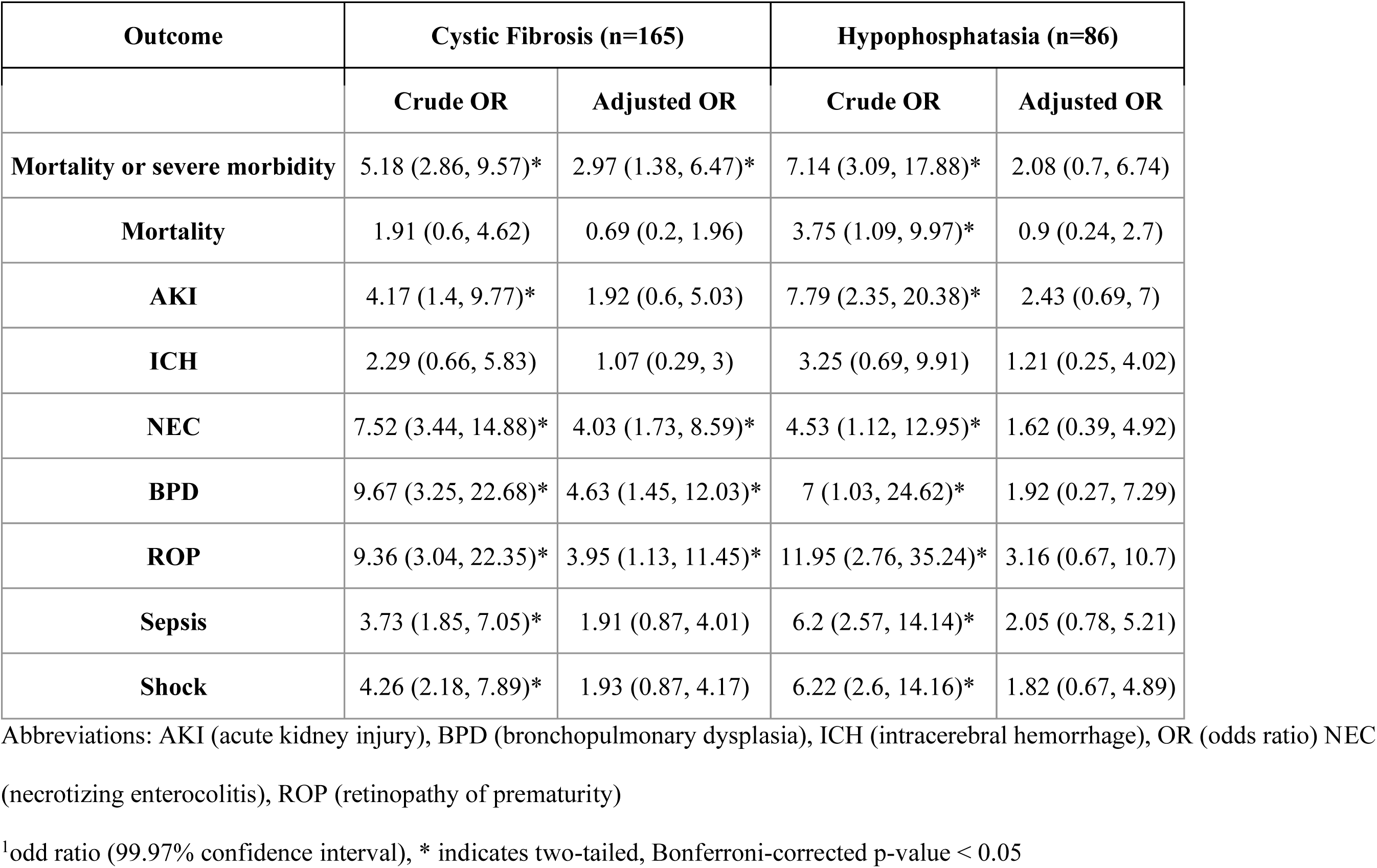
Odds of severe morbidity or mortality in preterm infants with miscellaneous genetic disorders^1^.

## Discussion

This study provides the most comprehensive determination to our knowledge of the prevalence of genetic disorders in preterm infants and short-term risk of morbidity and mortality conferred by genetic disorders in this population. We found that genetic disorders, consisting of aneuploidies, copy number variants, and monogenic disorders, were diagnosed in 1.3% of preterm infants. Trisomy 13, 18, and 21, and cystic fibrosis were associated with increased risk of morbidity or mortality after adjustment for known risk factors. Rates of comorbidities for preterm infants with trisomy 13 and 18 may be underestimated given mortality as a competing outcome, particularly following redirection to palliative goals of care.

The overall prevalence of genetic disorders may reflect a contribution of genetic disease to shorter gestation. Several of the most prevalent genetic disorders we analyzed are associated with preterm birth including hemoglobinopathies,^16^ cystic fibrosis,^17^ and common aneuploidies.^18^ This results from a complex interplay of factors including disrupted placentation,^18^ increased risk of fetal growth restriction and stillbirth, closer monitoring of pregnancy, greater maternal age contributing to aneuploidy and risk of preterm birth,^19^ as well as idiopathic factors.

We found prevalence rates of trisomy 13 and 18 lower than previously described in preterm infants,^12^ however our analysis used a gestational age-based cutoff rather than birth weight. Trisomy 13 and 18 cause fetal growth restriction. Given the same study population, a weight-based cutoff will select for infants with gestational age > 33 weeks with these genetic disorders and estimate a higher prevalence rate. Given these selection differences, it is difficult to compare morbidity and mortality between our cohorts and previous descriptions.^11^

Interestingly, we found increased adjusted odds of NEC, BPD, and ROP in preterm infants with cystic fibrosis. Infants with cystic fibrosis have been found to be born at earlier gestational age and with lower birth weight.^17,20^ However, after adjustment for these factors, we found an increased risk of morbidity. Intestinal disease caused by meconium peritonitis and meconium ileus may mimic or predispose infants to NEC.^21,22^ BPD and ROP may be attributable to disrupted lung disease, and exposure to oxygen.^23,24^ Maternal cystic fibrosis also contributes to neonatal morbidity,^25^ and may account for a portion of observed outcomes. However, maternal diagnoses were not available in our cohort.

This retrospective study has significant strengths and important weaknesses. It encompasses a large number of infants, cared for in multiple institutions, and is highly generalizable. The depth of data, including patient characteristics, diagnoses of genetic disorders, and co-morbidities should enable clinicians to better counsel families and inform clinical decision-making in rare presentations. It encompasses a broad period, during which genetic testing has undergone considerable change. Interestingly, we did not detect differences in the broad classes of genetic disorders diagnosed in preterm infants over time (**Figure 1**). However, our dataset lacked information regarding genetic testing modalities, and negative results. There may be underlying, dynamic changes that contributed to heterogeneity in the cohort with respect to year of birth. As genome-wide testing becomes more common, rare and uncommon genetic disorders may be diagnosed more frequently in this population.

This dataset lacks information regarding social determinants of health and maternal access to care (including genetic testing). Multiple factors could have contributed to over- or underestimation of the prevalence of genetic disorders. Preterm infants experience high rates of morbidity and mortality, which may prompt testing for genetic disease in some settings. Genetic disorders often present non-specifically in preterm infants,^4,26^ compared to more complete and well-known phenotypes found in term infants or pediatric or adult populations. Therefore, genetic disorders may have gone undetected in this cohort. Unfortunately, we do not have data regarding the extent of pre- and postnatal genetic testing, including negative results, in this cohort. At this time, broad, prospective screening for neonatal genetic disorders is not routine. Given the impact of genetic disorders on preterm morbidity and mortality in our study, there is a crucial need for broad, prospective studies to establish this. At present, a number of studies are underway to evaluate the potential of sequencing based newborn screening. In addition to determining the technical advantage of obtaining a molecular diagnosis of a limited number of rare biochemical disorders, this work may offer an opportunity to investigate the impact of a more diverse group of genetic disorders on neonatal care.

In summary, using a multicenter retrospective cohort, we found that genetic disorders are present in 1.3% of infants born before 34 weeks’ gestation, and that trisomy 13, 18, 21, and cystic fibrosis are associated with greater adjusted odds of death or severe morbidity. We determined the prevalence of 19 distinct genetic disorders, and the crude and adjusted risk of mortality and severe morbidities. These data may help clinicians provide more informed guidance to families and lead to better clinical decision-making in rare presentations.^27^ These data indicate the need for a greater index of suspicion for multiple critical illnesses, such as sepsis in the setting of trisomy 21 and NEC in the setting of cystic fibrosis. Given the trend towards greater adjusted odds of multiple morbid outcomes, clinicians should also consider genetic evaluation when caring for preterm infants with critical illnesses. Prospective, genome-wide screening is needed to determine the true prevalence of genetic disorders in this high-risk population, the contribution of genetic disorders to preterm birth, and the portion of disease in preterm infants attributable to genetic disorders.

## Data Availability

All data produced in the present study are available upon reasonable request to the authors.

## Acknowledgments

The authors wish to gratefully acknowledge Dr. Ali G. Gharavi (Columbia University Irving Medical Center) for their mentorship and thoughtful review of this project.

---

**Supplemental Table 1.**
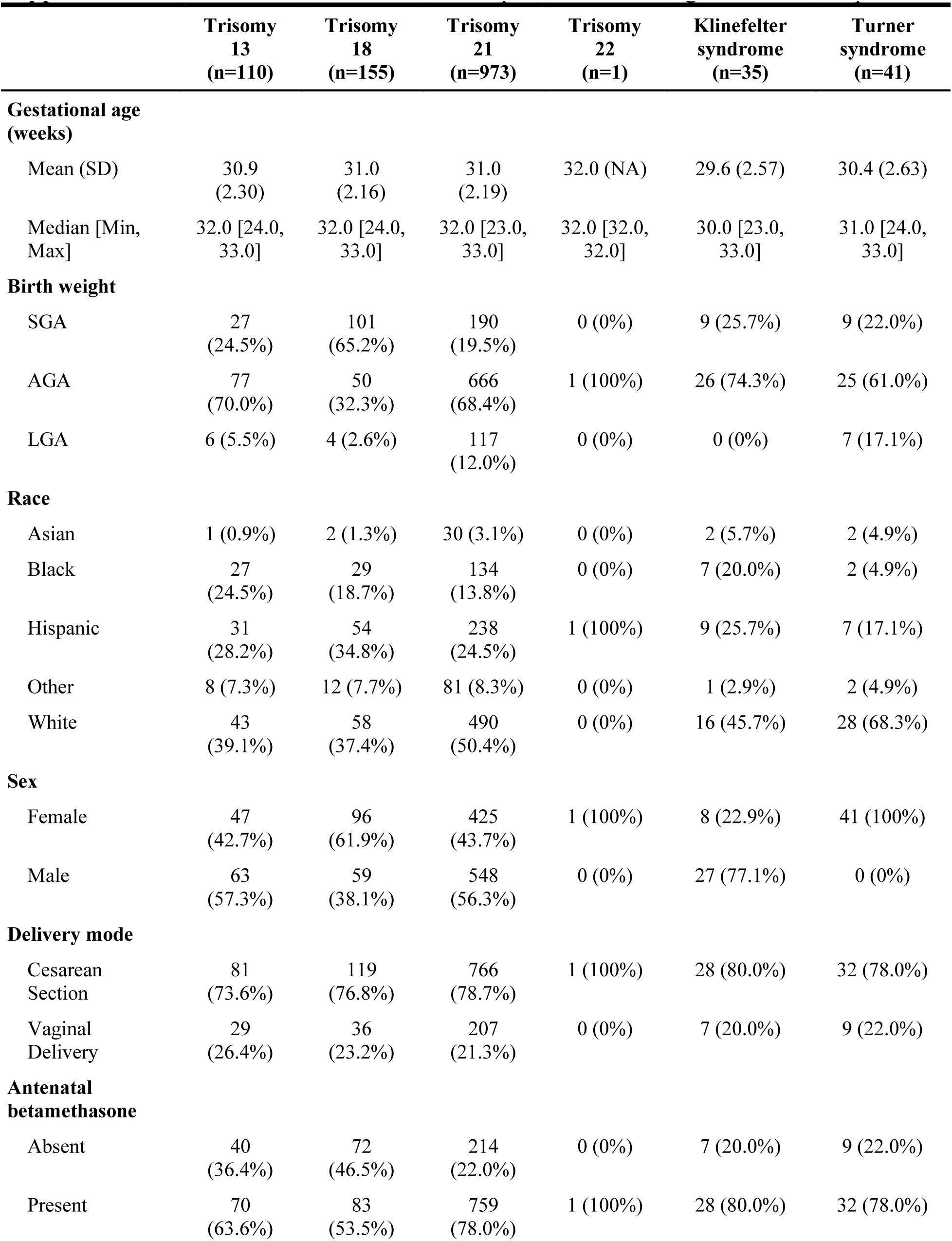
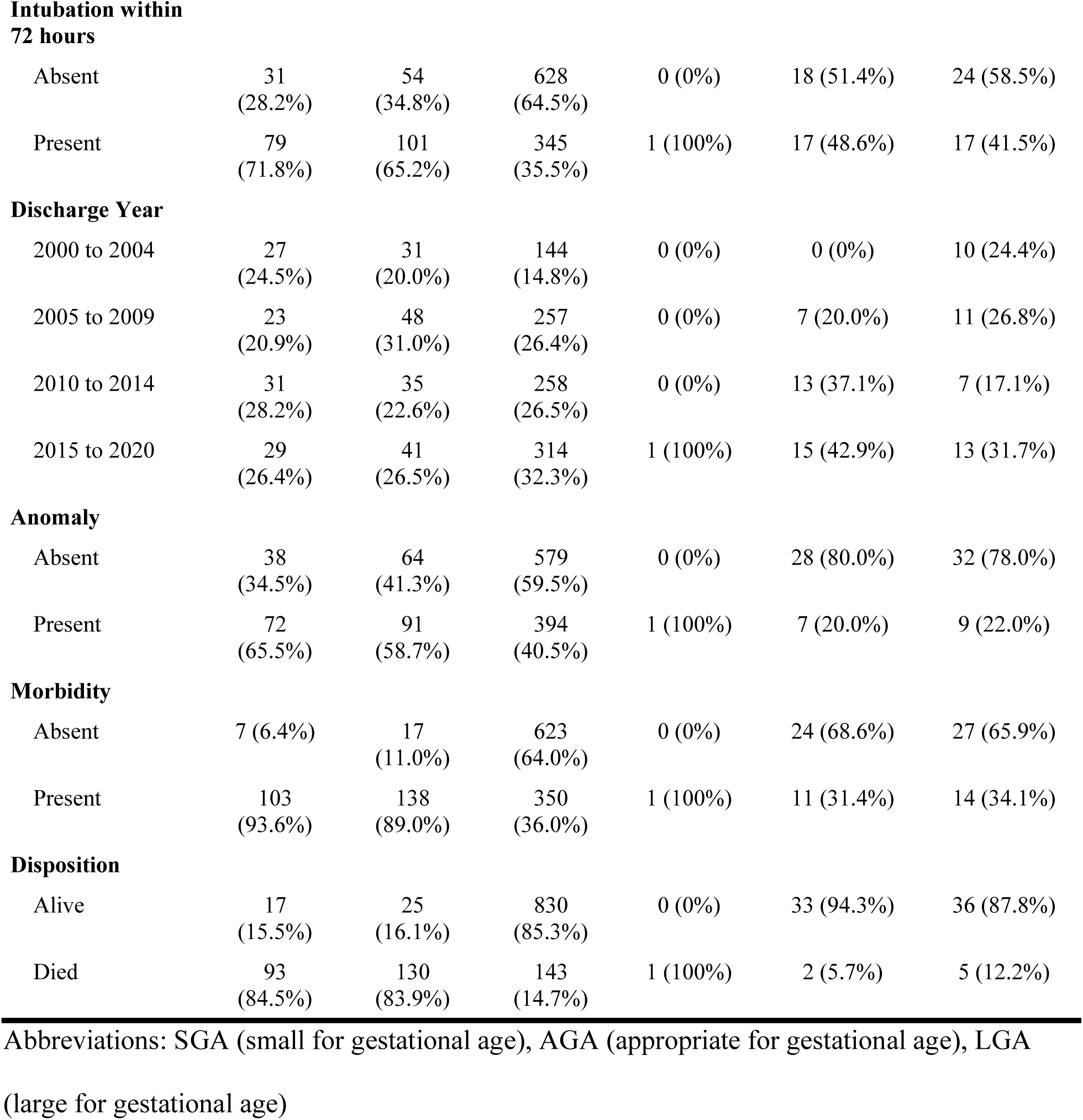
Baseline characteristics of preterm infants diagnosed with aneuploidies.

|  | **Trisomy**<br>**13**<br>**(n=110)** | **Trisomy**<br>**18**<br>**(n=155)** | **Trisomy**<br>**21**<br>**(n=973)** | **Trisomy**<br>**22**<br>**(n=1)** | **Klinefelter**<br>**syndrome**<br>**(n=35)** | **Turner**<br>**syndrome**<br>**(n=41)** |
| --- | --- | --- | --- | --- | --- | --- |
| **Gestational age**<br>**(weeks)** |  |  |  |  |  |  |
| Mean (SD) | 30.9<br>(2.30) | 31.0<br>(2.16) | 31.0<br>(2.19) | 32.0 (NA) | 29.6 (2.57) | 30.4 (2.63) |
| Median [Min,<br>Max] | 32.0 [24.0,<br>33.0] | 32.0 [24.0,<br>33.0] | 32.0 [23.0,<br>33.0] | 32.0 [32.0,<br>32.0] | 30.0 [23.0,<br>33.0] | 31.0 [24.0,<br>33.0] |
| **Birth weight** |  |  |  |  |  |  |
| SGA | 27<br>(24.5%) | 101<br>(65.2%) | 190<br>(19.5%) | 0 (0%) | 9 (25.7%) | 9 (22.0%) |
| AGA | 77<br>(70.0%) | 50<br>(32.3%) | 666<br>(68.4%) | 1 (100%) | 26 (74.3%) | 25 (61.0%) |
| LGA | 6 (5.5%) | 4 (2.6%) | 117<br>(12.0%) | 0 (0%) | 0 (0%) | 7 (17.1%) |
| **Race** |  |  |  |  |  |  |
| Asian | 1 (0.9%) | 2 (1.3%) | 30 (3.1%) | 0 (0%) | 2 (5.7%) | 2 (4.9%) |
| Black | 27<br>(24.5%) | 29<br>(18.7%) | 134<br>(13.8%) | 0 (0%) | 7 (20.0%) | 2 (4.9%) |
| Hispanic | 31<br>(28.2%) | 54<br>(34.8%) | 238<br>(24.5%) | 1 (100%) | 9 (25.7%) | 7 (17.1%) |
| Other | 8 (7.3%) | 12 (7.7%) | 81 (8.3%) | 0 (0%) | 1 (2.9%) | 2 (4.9%) |
| White | 43<br>(39.1%) | 58<br>(37.4%) | 490<br>(50.4%) | 0 (0%) | 16 (45.7%) | 28 (68.3%) |
| **Sex** |  |  |  |  |  |  |
| Female | 47<br>(42.7%) | 96<br>(61.9%) | 425<br>(43.7%) | 1 (100%) | 8 (22.9%) | 41 (100%) |
| Male | 63<br>(57.3%) | 59<br>(38.1%) | 548<br>(56.3%) | 0 (0%) | 27 (77.1%) | 0 (0%) |
| **Delivery mode** |  |  |  |  |  |  |
| Cesarean<br>Section | 81<br>(73.6%) | 119<br>(76.8%) | 766<br>(78.7%) | 1 (100%) | 28 (80.0%) | 32 (78.0%) |
| Vaginal<br>Delivery | 29<br>(26.4%) | 36<br>(23.2%) | 207<br>(21.3%) | 0 (0%) | 7 (20.0%) | 9 (22.0%) |
| **Antenatal**<br>**betamethasone** |  |  |  |  |  |  |
| Absent | 40<br>(36.4%) | 72<br>(46.5%) | 214<br>(22.0%) | 0 (0%) | 7 (20.0%) | 9 (22.0%) |
| Present | 70<br>(63.6%) | 83<br>(53.5%) | 759<br>(78.0%) | 1 (100%) | 28 (80.0%) | 32 (78.0%) |
| **Intubation within 72 hours** |  |  |  |  |  |  |
| Absent | 31<br>(28.2%) | 54<br>(34.8%) | 628<br>(64.5%) | 0 (0%) | 18 (51.4%) | 24 (58.5%) |
| Present | 79<br>(71.8%) | 101<br>(65.2%) | 345<br>(35.5%) | 1 (100%) | 17 (48.6%) | 17 (41.5%) |
| **Discharge Year** |  |  |  |  |  |  |
| 2000 to 2004 | 27<br>(24.5%) | 31<br>(20.0%) | 144<br>(14.8%) | 0 (0%) | 0 (0%) | 10 (24.4%) |
| 2005 to 2009 | 23<br>(20.9%) | 48<br>(31.0%) | 257<br>(26.4%) | 0 (0%) | 7 (20.0%) | 11 (26.8%) |
| 2010 to 2014 | 31<br>(28.2%) | 35<br>(22.6%) | 258<br>(26.5%) | 0 (0%) | 13 (37.1%) | 7 (17.1%) |
| 2015 to 2020 | 29<br>(26.4%) | 41<br>(26.5%) | 314<br>(32.3%) | 1 (100%) | 15 (42.9%) | 13 (31.7%) |
| **Anomaly** |  |  |  |  |  |  |
| Absent | 38<br>(34.5%) | 64<br>(41.3%) | 579<br>(59.5%) | 0 (0%) | 28 (80.0%) | 32 (78.0%) |
| Present | 72<br>(65.5%) | 91<br>(58.7%) | 394<br>(40.5%) | 1 (100%) | 7 (20.0%) | 9 (22.0%) |
| **Morbidity** |  |  |  |  |  |  |
| Absent | 7 (6.4%) | 17<br>(11.0%) | 623<br>(64.0%) | 0 (0%) | 24 (68.6%) | 27 (65.9%) |
| Present | 103<br>(93.6%) | 138<br>(89.0%) | 350<br>(36.0%) | 1 (100%) | 11 (31.4%) | 14 (34.1%) |
| **Disposition** |  |  |  |  |  |  |
| Alive | 17<br>(15.5%) | 25<br>(16.1%) | 830<br>(85.3%) | 0 (0%) | 33 (94.3%) | 36 (87.8%) |
| Died | 93<br>(84.5%) | 130<br>(83.9%) | 143<br>(14.7%) | 1 (100%) | 2 (5.7%) | 5 (12.2%) |

Abbreviations: SGA (small for gestational age), AGA (appropriate for gestational age), LGA (large for gestational age)

**Supplemental Table 2.** Baseline characteristics of preterm infants diagnosed with copy number variants.

|  | DiGeorge syndrome<br>(n=29) | Cri-du-chat syndrome<br>(n=16) | 13q-deletion syndrome<br>(n=7) |
| --- | --- | --- | --- |
| **Gestational age (weeks)** |  |  |  |
| Mean (SD) | 30.6 (2.40) | 31.3 (2.65) | 31.1 (1.35) |
| Median [Min, Max] | 31.0 [25.0, 33.0] | 32.0 [24.0, 33.0] | 31.0 [29.0, 33.0] |
| **Birth weight** |  |  |  |
| SGA | 3 (10.3%) | 2 (12.5%) | 3 (42.9%) |
| AGA | 23 (79.3%) | 14 (87.5%) | 4 (57.1%) |
| LGA | 3 (10.3%) | 0 (0%) | 0 (0%) |
| **Race** |  |  |  |
| Black | 9 (31.0%) | 6 (37.5%) | 4 (57.1%) |
| Hispanic | 9 (31.0%) | 4 (25.0%) | 3 (42.9%) |
| Other | 2 (6.9%) | 1 (6.3%) | 0 (0%) |
| White | 8 (27.6%) | 5 (31.3%) | 0 (0%) |
| Asian | 1 (3.4%) | 0 (0%) | 0 (0%) |
| **Sex** |  |  |  |
| Female | 17 (58.6%) | 12 (75.0%) | 3 (42.9%) |
| Male | 12 (41.4%) | 4 (25.0%) | 4 (57.1%) |
| **Delivery mode** |  |  |  |
| Cesarean Section | 24 (82.8%) | 12 (75.0%) | 5 (71.4%) |
| Vaginal Delivery | 5 (17.2%) | 4 (25.0%) | 2 (28.6%) |
| **Antenatal betamethasone** |  |  |  |
| Absent | 9 (31.0%) | 1 (6.3%) | 2 (28.6%) |
| Present | 20 (69.0%) | 15 (93.8%) | 5 (71.4%) |
| **Intubation within 72 hours** |  |  |  |
| Absent | 14 (48.3%) | 8 (50.0%) | 4 (57.1%) |
| Present | 15 (51.7%) | 8 (50.0%) | 3 (42.9%) |
| **Discharge Year** |  |  |  |
| 2000 to 2004 | 1 (3.4%) | 2 (12.5%) | 0 (0%) |
| 2005 to 2009 | 8 (27.6%) | 5 (31.3%) | 0 (0%) |
| 2010 to 2014 | 6 (20.7%) | 2 (12.5%) | 1 (14.3%) |
| 2015 to 2020 | 14 (48.3%) | 7 (43.8%) | 6 (85.7%) |
| **Anomaly** |  |  |  |
| Absent | 13 (44.8%) | 8 (50.0%) | 5 (71.4%) |
| Present | 16 (55.2%) | 8 (50.0%) | 2 (28.6%) |
| **Morbidity** |  |  |  |
| Absent | 17 (58.6%) | 11 (68.8%) | 5 (71.4%) |
| Present | 12 (41.4%) | 5 (31.3%) | 2 (28.6%) |
| **Disposition** |  |  |  |
| Alive | 25 (86.2%) | 14 (87.5%) | 6 (85.7%) |
| Died | 4 (13.8%) | 2 (12.5%) | 1 (14.3%) |

Abbreviations: SGA (small for gestational age), AGA (appropriate for gestational age), LGA
(large for gestational age)

**Supplemental Table 3.**
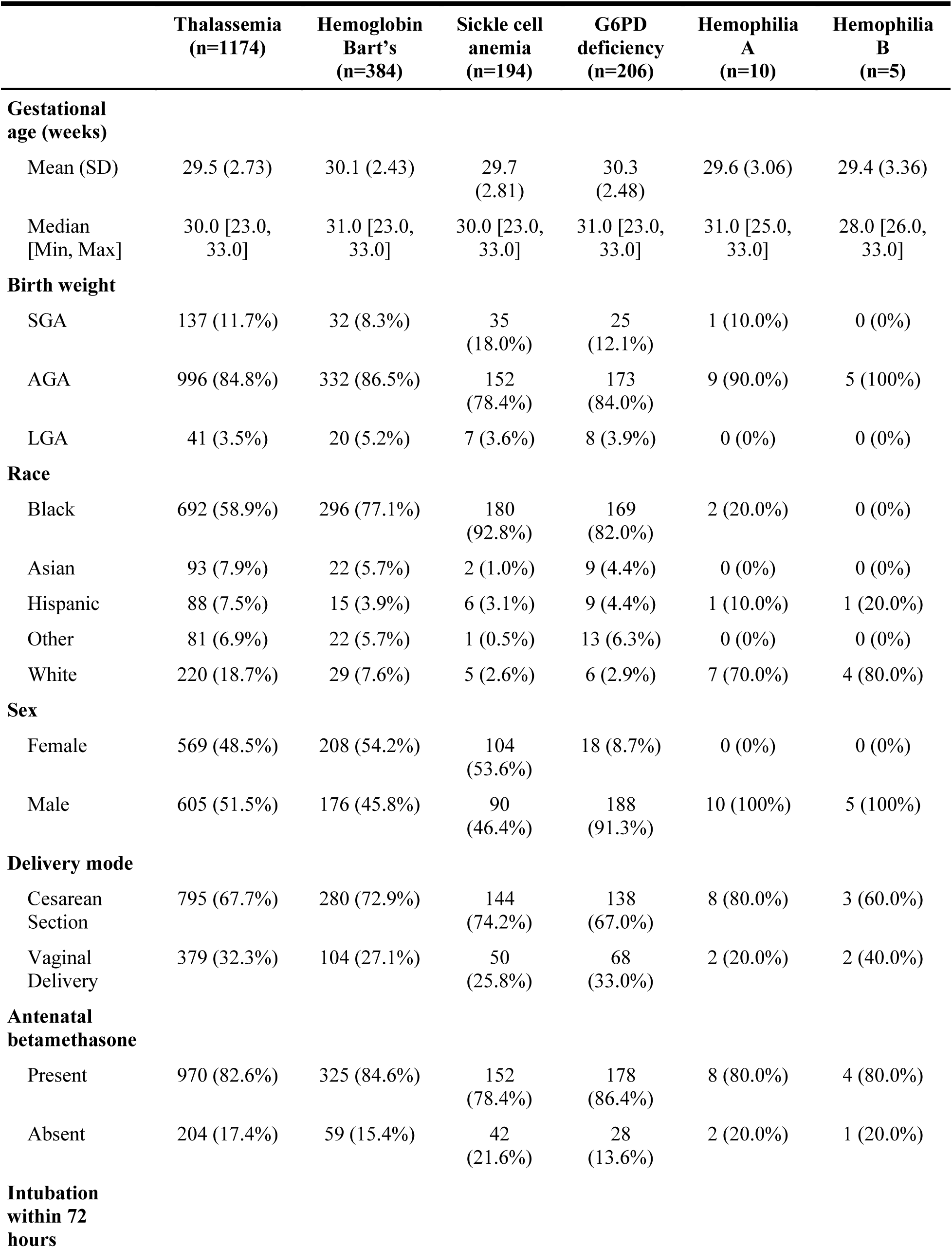
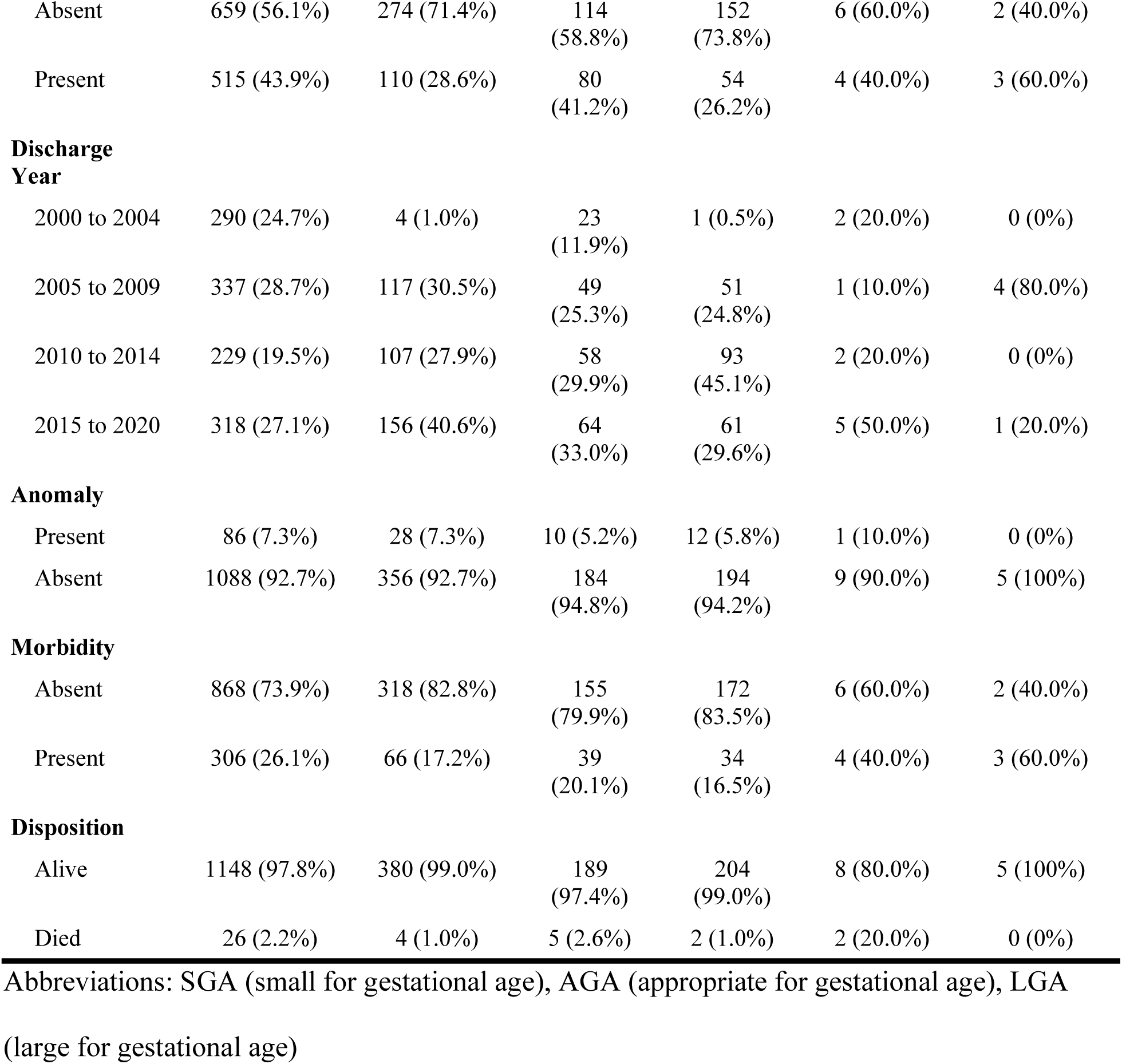
Baseline characteristics of preterm infants diagnosed with hematologic disorders.

**Supplemental Table 4.** Baseline characteristics of preterm infants diagnosed with miscellaneous genetic disorders.

|  | **Cystic**<br>**fibrosis**<br>**(n=165)** | **Hypophosphatasia**<br>**(n=86)** | **Fragile X**<br>**syndrome**<br>**(n=7)** | **Fanconi**<br>**anemia**<br>**(n=2)** |
| --- | --- | --- | --- | --- |
| **Gestational age**<br>**(weeks)** |  |  |  |  |
| Mean (SD) | 29.0 (3.12) | 27.6 (2.84) | 30.1 (2.34) | 29.5 (4.95) |
| Median [Min,<br>Max] | 29.0 [23.0,<br>33.0] | 27.0 [23.0, 33.0] | 31.0 [26.0,<br>32.0] | 29.5 [26.0,<br>33.0] |
| **Birth weight** |  |  |  |  |
| SGA | 26 (15.8%) | 32 (37.2%) | 0 (0%) | 2 (100%) |
| AGA | 126<br>(76.4%) | 52 (60.5%) | 5 (71.4%) | 0 (0%) |
| LGA | 13 (7.9%) | 2 (2.3%) | 2 (28.6%) | 0 (0%) |
| **Race** |  |  |  |  |
| Asian | 1 (0.6%) | 4 (4.7%) | 0 (0%) | 2 (100%) |
| Black | 17 (10.3%) | 21 (24.4%) | 1 (14.3%) | 0 (0%) |
| Hispanic | 14 (8.5%) | 22 (25.6%) | 0 (0%) | 0 (0%) |
| Other | 6 (3.6%) | 6 (7.0%) | 0 (0%) | 0 (0%) |
| White | 127<br>(77.0%) | 33 (38.4%) | 6 (85.7%) | 0 (0%) |
| **Sex** |  |  |  |  |
| Female | 71 (43.0%) | 40 (46.5%) | 2 (28.6%) | 2 (100%) |
| Male | 94 (57.0%) | 46 (53.5%) | 5 (71.4%) | 0 (0%) |
| **Delivery mode** |  |  |  |  |
| Cesarean Section | 115<br>(69.7%) | 69 (80.2%) | 5 (71.4%) | 2 (100%) |
| Vaginal Delivery | 50 (30.3%) | 17 (19.8%) | 2 (28.6%) | 0 (0%) |
| **Antenatal**<br>**betamethasone** |  |  |  |  |
| Absent | 33 (20.0%) | 18 (20.9%) | 2 (28.6%) | 2 (100%) |
| Present | 132<br>(80.0%) | 68 (79.1%) | 5 (71.4%) | 0 (0%) |
| **Intubation within**<br>**72 hours** |  |  |  |  |
| Absent | 52 (31.5%) | 21 (24.4%) | 3 (42.9%) | 1 (50.0%) |
| Present | 113<br>(68.5%) | 65 (75.6%) | 4 (57.1%) | 1 (50.0%) |
| **Discharge Year** |  |  |  |  |
| 2000 to 2004 | 68 (41.2%) | 27 (31.4%) | 2 (28.6%) | 0 (0%) |
| 2005 to 2009 | 56 (33.9%) | 26 (30.2%) | 0 (0%) | 0 (0%) |
| 2010 to 2014 | 24 (14.5%) | 19 (22.1%) | 1 (14.3%) | 1 (50.0%) |
| 2015 to 2020 | 17 (10.3%) | 14 (16.3%) | 4 (57.1%) | 1 (50.0%) |
| **Anomaly** |  |  |  |  |
| Absent | 126<br>(76.4%) | 78 (90.7%) | 5 (71.4%) | 2 (100%) |
| Present | 39 (23.6%) | 8 (9.3%) | 2 (28.6%) | 0 (0%) |
| **Morbidity** |  |  |  |  |
| Absent | 68 (41.2%) | 29 (33.7%) | 6 (85.7%) | 1 (50.0%) |
| Present | 97 (58.8%) | 57 (66.3%) | 1 (14.3%) | 1 (50.0%) |
| **Disposition** |  |  |  |  |
| Alive | 149<br>(90.3%) | 71 (82.6%) | 7 (100%) | 1 (50.0%) |
| Died | 16 (9.7%) | 15 (17.4%) | 0 (0%) | 1 (50.0%) |

SGA: small for gestational age; AGA: appropriate for gestational age; LGA: large for gestational age.

**Supplemental Table 5.** Instances of severe morbidity and mortality in preterm infants with genetic disorders.

| Genetic Disorder | Mortality or Any Severe Morbidity | Mortality | AKI | ICH | NEC | BPD | ROP | Sepsis | Shock |
| --- | --- | --- | --- | --- | --- | --- | --- | --- | --- |
| *No genetic diagnosis* | 68326 | 16861 | 9021 | 12289 | 10933 | 3953 | 3836 | 28874 | 30087 |
| *Any genetic diagnosis* | 1526 | 565 | 279 | 204 | 232 | 172 | 72 | 585 | 615 |
| Aneuploidy | 883 | 480 | 171 | 93 | 101 | 105 | 16 | 258 | 341 |
| Trisomy 13 | 103 | 93 | 18 | 0 | 5 | 7 | 1 | 9 | 20 |
| Trisomy 18 | 138 | 130 | 13 | 2 | 5 | 10 | 0 | 11 | 31 |
| Trisomy 21 | 350 | 143 | 88 | 41 | 51 | 48 | 4 | 127 | 155 |
| Trisomy 22 | 1 | 1 | 1 | 0 | 0 | 0 | 0 | 0 | 0 |
| Klinefelter syndrome | 11 | 2 | 2 | 2 | 0 | 1 | 0 | 8 | 4 |
| Turner syndrome | 14 | 5 | 5 | 3 | 2 | 1 | 0 | 9 | 6 |
| Unspecified Aneuploidy | 266 | 106 | 44 | 45 | 38 | 38 | 11 | 94 | 125 |
| Copy number variation disorder | 35 | 14 | 7 | 1 | 8 | 5 | 0 | 16 | 13 |
| 13q deletion syndrome | 2 | 1 | 0 | 1 | 0 | 0 | 0 | 0 | 0 |
| Cri-du-chat syndrome | 5 | 2 | 2 | 0 | 2 | 1 | 0 | 4 | 2 |
| DiGeorge syndrome | 12 | 4 | 1 | 0 | 1 | 2 | 0 | 6 | 4 |
| Unspecified CNV | 16 | 7 | 4 | 0 | 5 | 2 | 0 | 6 | 7 |
| Monogenic disorder | 608 | 71 | 101 | 110 | 123 | 62 | 56 | 311 | 261 |
| Hematologic disorder | 452 | 39 | 66 | 85 | 76 | 36 | 28 | 232 | 174 |
| Thalassemia | 306 | 26 | 47 | 61 | 55 | 20 | 25 | 156 | 125 |
| Hemoglobin bart's | 66 | 4 | 4 | 8 | 9 | 5 | 0 | 35 | 25 |
| Sickle cell disease | 39 | 5 | 9 | 6 | 2 | 5 | 2 | 21 | 12 |
| G6PD deficiency | 34 | 2 | 5 | 7 | 10 | 5 | 1 | 15 | 9 |
| Hemophilia A | 4 | 2 | 1 | 2 | 0 | 1 | 0 | 3 | 2 |
| Hemophilia B | 3 | 0 | 0 | 1 | 0 | 0 | 0 | 2 | 1 |
| Miscellaneous | 156 | 32 | 35 | 25 | 47 | 26 | 28 | 79 | 87 |
| Cystic fibrosis | 97 | 16 | 18 | 14 | 35 | 18 | 17 | 45 | 51 |
| Fragile X syndrome | 1 | 0 | 0 | 0 | 0 | 0 | 0 | 1 | 1 |
| Hypophosphatasia | 57 | 15 | 16 | 10 | 12 | 7 | 11 | 33 | 34 |
| Fanconi Anemia | 1 | 1 | 1 | 1 | 0 | 1 | 0 | 0 | 1 |
| Multiple Disorders | 6 | 1 | 2 | 1 | 3 | 2 | 1 | 5 | 2 |

Abbreviations: AKI (acute kidney injury), BPD (bronchopulmonary dysplasia), ICH (intracerebral hemorrhage), OR (odds ratio) NEC (necrotizing enterocolitis), ROP (retinopathy of prematurity)

